# Facilitating Healing for Black Women Experiencing Gendered Racism and Traumatic Stress: The Moderation of Psychosocial Resources

**DOI:** 10.1101/2023.04.17.23288699

**Authors:** Tiffany R. Williams, Christy L. Erving, Whitney Frierson, Fanchen Gao, Jeffery E Bass, Reniece Martin, Taeja Mitchell

## Abstract

Black women must navigate a tumultuous sociopolitical terrain while simultaneously managing their psychological health. Experiences of gendered racism increase Black women’s vulnerability to psychological distress. Gendered racial microaggressions, a specific type of microaggression, account for the intricate ways racism and sexism intersect. The association between Black women’s experiences of gendered racial microaggressions and traumatic stress was investigated among 201 Black female-identified undergraduate and graduate students attending a Historically Black College or University. Whether psychosocial resources (i.e., resilience, social support, mastery, self-esteem) moderated the linkage between gendered racial microaggressions and traumatic stress was also examined. Gendered racial microaggressions were positively associated with traumatic stress. The microaggression Assumptions of Beauty and Sexual Objectification was the most strongly associated with traumatic stress, followed by Angry Black Woman. Resilience and mastery were protective factors, reducing the influence of gendered racial microaggressions on traumatic stress. In addition, high levels of social support reduced the impact of Assumptions of Beauty and Sexual Objectification on traumatic stress. To foster healing and posttraumatic growth for Black women, psychologists must decolonize their understanding and treatment of mental illness. Practice and research implications are discussed.

---

The United States (U.S.) climate, rooted in Eurocentric ideologies, systems, and institutions, perpetuates hate, violence, and oppression toward Black women. Informed by sexism, racism, and other forms of discrimination, Black women must tirelessly navigate the tumultuous sociopolitical terrain, while simultaneously managing their psychological health. Neglected, overlooked, and invisible are Black women’s experiences in mainstream media or dominant culture, despite the complexity, intensity, and profound effects on their well-being (DuMonthier et al., 2020). Black women’s experiences are complex and unique, and at times, make them more vulnerable to traumatic stress than their White counterparts. In fact, a national study reported that Black women are at 34% greater risk for experiencing PTSD in their lifetime relative to their White female counterparts (Erving et al., 2019). For example, Black women are reported to experience disproportionate and highest rates of intimate partner violence, sexual assault, and childhood sexual abuse (DuMonthier et al., 2020; Green, 2017). As compared to White women, four out of 10 Black women report experiences of physical violence; they are 3.5 times more likely to be killed by men and are more likely to experience homicide by a significant other, spouse, or family member (Violence Policy Center, 2020).

Whether at home or in the community, navigating intersecting oppressions contributes to psychological distress and emotional pain (Jones & Shorter-Gooden, 2003; Lewis & Neville, 2015). Violence and hate are the products of intersecting experiences of gender- and race-based discrimination, also known as gendered racism, against Black women. Not only as direct recipients but also as indirect recipients, emotional anguish and undue stress can proliferate, as they are inundated with a barrage of videos and photos on social media or news outlets exhibiting gendered racism and violence toward Black women (DuMonthier et al., 2020; Meshberg-Cohen et al., 2016). A natural direct result of managing gendered racism is trauma-related reactions, which increase Black women’s susceptibility to psychological distress (Green, 2017; Meshberg-Cohen et al., 2016). Case in point, Black women exponentially experience higher rates of trauma as compared to other racial groups; nearly all Black women (roughly 94.2%) are reported to have experienced at least one trauma that meets the criteria for posttraumatic stress disorder in their lifetime (Meshberg-Cohen et al., 2016).

Previous research demonstrates how gendered racism stems from racialized stereotypes that permeate the dominant discourse, mainstream culture, and societal attitudes about Black women (Cheers, 2020; West, 2018; Woods-Giscombé et al., 2016). Gendered racial microaggressions, an expansion of Sue and colleagues’ microaggressions taxonomy (2007), underlie gendered racism and highlight the subtle, cumulative, and nuanced slights while accounting for the intersectional experience of racism and sexism for Black women (Lewis & Neville, 2015). Experiences of gendered racial microaggressions are linked with poor mental health (Castelin & White, 2022; Williams & Lewis, 2019; Wright & Lewis, 2020).

Independently, resilience, social support, mastery, and self-esteem are identified as protective factors, or psychosocial resources, known to improve subjective well-being for Black women (Constantine et al., 2002; Keith et al., 2010; Mushonga et al., 2021). However, the effects of these psychosocial resources on Black women’s experience of gendered racial microaggressions or traumatic stress are severely understudied. To date, there are only two other studies that investigate the relationship between gendered racial microaggressions and traumatic stress (Dale & Safren, 2019; Moody & Lewis, 2019). Both demonstrate a positive relationship exists, yet none of these studies investigated the impacts of psychosocial resources. Liberation psychology guides our purpose for the present study. Moving toward a liberation and decolonization assessment and intervention to facilitate healing for Black women is necessary to avoid further oppressing them (Tummala-Narra, 2007; Comas-Díaz, 2016). We expect that capitalizing on Black women’s psychosocial resources will reduce the effects of gendered racism on traumatic stress. The purpose of this research was to examine the association between gendered racial microaggressions and traumatic stress and to determine if psychosocial resources mitigate the negative effects of gendered racism on Black women’s reports of trauma symptoms.

## Gendered Racism

Essed (1991) defines gendered racism as a concept reflecting Black women’s intersectional experiences of racism and sexism, symbolizing multiple forms of oppression. These intersectional experiences increase Black women’s vulnerability to mental health conditions and are significantly linked with psychological distress (Williams & Lewis, 2019). Racialized stereotypes inform gendered racism and reveal how they are embedded in U.S. mainstream culture and societal messaging to perpetuate the oppression of Black women (Jones & Shorter-Gooden, 2003). Racial stereotyping is characterized as a method of defining and reifying cultural differences (Pickering, 2004); moreover, some racialized stereotypes are also gendered. The most prevalent racialized stereotypes that misrepresent Black women are Sapphire/Angry Black woman, Welfare Queen, Jezebel, Mammy, and Superwoman/Strong Black woman (Cheers, 2020; West, 2018; Woods-Giscombé et al., 2016). Through repeated exposure to the portrayal of racialized and oppressive representation, people internalize these misrepresentations building schemas and false narratives about Black women and their experiences (Lewis & Neville, 2015; Jones & Shorter-Gooden, 2003; West, 2018).

## Gendered Racial Microaggressions

Sue and colleagues (2007) describe microaggressions as everyday slights and insults deriving from racial assumptions about criminality, intelligence, cultural values, and citizenship, and minimization or denial of people of color’s racialized experiences. Microaggressions are experienced based on the intersection of socially oppressed identities. Gendered racial microaggressions (GRMS) are a specific type of microaggression that account for the intricate ways that racism intersects with sexism. GRMS reflect racial and gender bias and are described with four assumptions: (a) beauty and sexual objectification, (b) silenced and marginalized, (c) strong Black women stereotype, and (d) angry Black women stereotype (Lewis & Neville, 2015). Black women learn to manage internalized messages that stem from their experiences of GRMS, such as powerlessness, invisibility, and beliefs about their sexuality, attractiveness, or physical appearance (e.g., hairstyles, body size) (Lewis et al., 2016). Exposure to gendered racism and racialized stereotypes are quite insidious as Black women are inundated with messaging in media, news outlets, places of employment, and other spaces that they occupy (Cheers, 2020; Jones & Shorter-Gooden, 2003; West, 2018). The cumulative and perpetual management of GRMS contributes to detrimental effects on Black women’s mental health.

## Effects on Mental Health

An abundance of research demonstrates the detrimental impact of racial stress on Black women’s psychological and physical health (Chinn et al. 2021; DuMonthier et al., 2020). Aside from navigating gendered racism, Black women are already at a particularly high risk of experiencing at least one trauma in their lifetime partly due to higher exposure to violence, vulnerability to homicide, and increased susceptibility to emotional, physical, and sexual abuse (DuMonthier et al., 2020; Cimino et al. 2019). There is evidence illustrating the effects of gendered and racial microaggressions on Black women’s mental health. For example, gendered racism is associated with increased symptoms of depression (Erving et al., 2022; Williams & Lewis, 2019), anxiety (Wright & Lewis, 2020), trauma (Dale & Safran, 2019; Moody & Lewis, 2019), suicidal behaviors, (Castelin & White, 2022), and body image (Winter et al., 2019).

In addition, specific dimensions of GRMS may be more trauma-inducing than others, as a recent study showed that Black women being perceived as angry was associated with higher depressive symptoms to a greater degree than other gendered racial microaggressions. In a study focused on the linkage between GRMS and PTSD among Black women living with HIV, Assumptions of Beauty and Sexual Objectification were associated with a higher risk of traumatic symptoms, but Strong Black Woman stereotypes were inversely associated with aspects of trauma (Dale & Safren, 2019). That is, Black women reporting Strong Black woman stereotypes have a lower risk of PTSD. These results suggest that certain gendered racial microaggressions may elicit traumatic stress while other dimensions may decrease it among Black women. Less is known about psychosocial mechanisms that reduce the impact of gendered racism on Black women’s experiences of traumatic stress. Psychosocial factors that reduce the effects of gendered racism must be identified to promote healing for Black women.

## Psychosocial Resources

Psychological distress, healing, and wellness are concepts rooted in Eurocentric values, assumptions, and ideals (Quiñones-Rosado, 2020). To foster healing for Black women, it is essential that psychologists decolonize their understanding and treatment of mental illness. Redirection and focus on identifying one’s personal and collective strengths and internal resources, informed by their culture, will help Black women embrace adversity and overcome hardship (Tummala-Narra, 2007; Quiñones-Rosado, 2020). Healing is facilitated through restorative practices. Restoration and balance are obtained through “physical, mental, spiritual, and emotional” components of the self, yet within the context of one’s community (Quiñones-Rosado, 2020, pp. 54). Integral to the process of healing and restoration is engaging in restorative activities that sustain balance or harmony. Harmony is characterized as the “intuitive directing of attention, awareness, consciousness, and wisdom” within the context of one’s ascribed cultural values and beliefs (Quiñones-Rosado, 2020, pp. 54). Engaging in restorative practices requires consciousness, intentionality, and congruence with one’s customs, culture, and worldviews (Tummala-Narra, 2007; Quiñones-Rosado, 2020). Identifying Black women’s psychosocial resources is key in helping them achieve balance and harmony. Equally important is facilitating critical consciousness of systemic oppressions (e.g., gendered racism) that disrupt healing, exacerbate distress, and generate disharmony and imbalance (Quiñones-Rosado, 2020). Here, we engage the possibility that psychosocial resources are restorative and healing practices that enable Black women to resist GRMS. Four psychosocial resources are discussed below.

Resilience is described as the intrapersonal process of overcoming adversity and “bouncing back” from hardship with the capacity to continue day-to-day functioning (Connor & Davidson, 2003). The complexity of being a Black woman contributes to the acquired level of resilience they have developed, which is necessary to thrive. Black women’s resilience comprises reframing stressors, identifying and utilizing resources, embracing optimism, and asserting boundaries through reducing problematic social interactions (Mushonga et al., 2021). Resilience is known to reduce psychological distress, and suicidal behaviors among Black women (Castelin & White, 2022; Spates & Slatton, 2021). For example, drawing on interviews with Black women, Spates and Slatton (2021) report that Black women utilize a “repertoire of resilience” that includes years of strength building, a collective experience of struggle, and a positive self-evaluation challenging dominant narratives about privilege. This repertoire motivates Black women to remain resilient during difficult times. Furthermore, social support, religion, and spirituality contribute to higher levels of resilience. Resilience facilitates posttraumatic growth among Black women; specifically, spirituality, motherhood, and gratitude contributed to resilience development (Mushonga et al., 2021). Research examining the connection between resilience and key components of Black culture suggested racial socialization and cultural support networks also contribute to resilience development (Brown, 2008). In an investigation on the correlation between racial socialization messages and perceived social support, Brown (2008) found that cultural factors, messages that stressed cultural pride, and knowledge of one’s African ancestry promoted resilience.

Social support is identified as a culturally concrete form of coping to manage adversity and hardships for Black women (Constantine et al., 2002). Traditional examples of support networks include immediate and extended family, the church, fictive kin, and the community (Brown, 2008; Taylor et al., 2001). Friends are another key source of social support for Black women, providing social companionship, emotional support, and practical assistance (Gray & Keith, 2001). In general, these varied sources of social support often contribute to Black women’s mental health by alleviating the impact of stressful events (Erving et al., 2022; Gray & Keith, 2001). Nonetheless, social support is often recognized as beneficial for Black women’s mental health (Sloan et al., 1996). Historically, churches have been integral in providing support networks, spaces for healing, communication, and inspiration, and opportunities for socialization and training development (Brown, 2008; Patton & McClure, 2009). In sum, research demonstrates the effectiveness of using a support system in coping with gender racism and psychological distress for Black women (Lewis et al., 2012; Neal-Barnett et al., 2011).

Personal mastery is a psychological concept that broadly refers to a person’s beliefs about one’s individual agency, locus of control, and own sense of self (Pearlin & Schooler, 1978; Keith et al., 2010). Mastery has been researched in connection with forms of trauma and it is associated with resilience, coping, and positive mental and physical health outcomes (Glister, 2016; Keith et al., 2010; Zanbar et al., 2021). People with a high sense of mastery use this resource as a coping mechanism by assessing their stressors and determining harm which aids in the likelihood of overcoming them (Glister, 2016; Pearlin & Schooler, 1978). Mastery is a pertinent factor in understanding how undue stress can be mitigated when discrimination negatively impacts Black women (Keith et al., 2010). Research demonstrates mastery plays a role in partially mitigating Black women’s psychological distress (Erving et al., 2021). For example, Erving et al. (2022) showed that mastery helped to reduce the impact of gendered racial microaggressions on depressive symptoms by 39.62%. One reason that mastery may serve as a protective psychosocial resource for Black women experiencing gendered racism is that high levels of mastery can influence Black women to remain strong despite discriminatory experiences.

Self-esteem is described as a person’s attitudes, feelings, and perceptions of oneself and the evaluation and judgments of these factors in relation to oneself (Rosenburg, 1979). Research on self-esteem suggests it plays a significant role in Black women’s psychological health (Erving et al., 2021; Watson et al., 2016). On the one hand, Black women’s experiences of intersecting oppressions can deteriorate their self-esteem and exacerbate psychological distress (Defrancisco & Chatham-Carpenter, 2000). On the other hand, self-esteem is a protective factor that reduces the effects of racist experiences and psychological distress when in conjunction with other factors, such as mastery and social support (Erving et al., 2021; Moody & Lewis, 2019). Self-esteem is a construct that partially reduces the effects of racist- and trauma-related experiences among women of color (Watson et al., 2016). Gendered racism has a profound yet harmful impact on Back women. Alone these psychosocial resources may be insufficient to reduce the full effects of gendered racism but collectively they have the potential to improve Black women’s healing and reduce the effects of traumatic stress.

## The Current Study

### Black women continue to face gendered racism and suffer the deleterious effects

Psychologists must use culturally sensitive treatments, even drawing upon Black women’s own internal resources, to endure, cope, and heal from these experiences (Castelin & White, 2022; Erving et al., 2020; Mushonga et al., 2021) Liberation psychology and decolonizing our perspectives on healing and wellness will challenge us to think critically about Black women’s intersectional experiences, conceptualize these experiences in the context of their culture and environment, and capitalize on their psychosocial resources to promote balance and harmony (Comas-Díaz, 2016; Quiñones-Rosado, 2020). We argue that the facilitation of healing for Black women requires an intersectional process comprising multiple psychosocial resources. Moreover, it is important to identify psychosocial resources that reduce the effects of gendered racism. We add to the extant literature proposing that psychosocial resources will reduce, but not entirely eliminate, the effects of gendered racism on traumatic stress. We hypothesize:

1. Greater frequency of reporting gendered and racial microaggressions will be positively associated with traumatic stress. We build on prior research by assessing which dimensions of gendered racial microaggressions are associated with higher traumatic stress.
2. Greater endorsement of gendered and racial microaggressions will be associated with lower levels of psychosocial resources (i.e., resilience, social support, mastery, and self-esteem).
3. High levels of psychosocial resources (i.e., resilience, social support, mastery, and self-esteem) will be associated with lower levels of traumatic stress.
4. Psychosocial resources will moderate the relationship between gendered racial microaggressions and traumatic stress, such that the association between gendered racial microaggressions and traumatic stress will be weaker among Black women with high levels of each psychosocial resource.

## Method

### Participants and Procedure

Participants in this study were a part of a larger mixed-methods study, the *Gendered Racism and Well-Being Questionnaire*, which consisted of 234 Black female-identified undergraduate and graduate students attending a southern Historically Black College or University (HBCU). The ages of the participants ranged from 18 to 65 years old. About 43% identified as first-generation college students. A broad range of degree programs and majors were reported (e.g., Psychology, Health Sciences, Nursing, and Chemistry). In terms of sexual identity, 84% identified as heterosexual, 3% as lesbian, 11% as bisexual, and 2% as other. Relationship status indicated that most participants were single (75%), while the remaining were either married (14%), or other (11%). In terms of children, 76% had no children, 9% had one child, 8% had two children, and 7% had three or more children. For employment status, participants reported as full-time (36%), part-time (34%,), not employed (27%), or other (3%).

Prior to recruitment, the Institutional Review Board approved the study. Participants completed an online survey via Qualtrics (2022) between September 2020 and April 2021. Participant recruitment was conducted through two phases. First, students enrolled in psychology courses were recruited via Sona Systems and received extra credit for participation. Second, through an email listserv, students throughout the university received recruitment fliers with a link to the Qualtrics survey. Each participant entered a raffle for the chance to win one of six $25 Amazon Gift Cards as compensation for their time. Participants were provided mental health resources at the beginning and end of the survey. The survey took about 30-40 minutes to complete. We report all information about data exclusions, sample size, management of missing data, and measures. Data is available upon request.

## Measures

### Gendered Racism

The Gendered Racial Microaggressions Scale (GRMS; Lewis & Neville, 2015) is a 26-item measure used to assess Black women’s experiences of everyday and subtle gendered racial microaggressions. Participants were asked to indicate the frequency of their experiences within the past year on a 5-point Likert-type scale ranging from 0 (n*ever*) to 4 (a *few times a month or more*). The average scores are then used to calculate a total mean frequency score. GRMS has four subscales including Assumptions of Beauty and Sexual Objectification (e.g., “Someone made negative comments about my hair when natural”; α = .92); Silenced and Marginalized (e.g., “I have been disrespected in the workplace”; α = .88); Strong Black Woman Stereotype (e.g., “I have been told that I am too assertive”; α = .75), and Angry Black Woman Stereotype (3-items; e.g., “Someone accused me of being angry when speaking calm”; α =.80). High internal consistency is reported on the total measure α = .92 (Lewis & Neville, 2015) and α = .93 (Moody & Lewis, 2019; Wright & Lewis, 2020). Internal reliability for the current study was very high (α = .95). Convergent validity was supported through significant positive correlations with similar measures of microaggressions and sexist events (Lewis & Neville, 2015).

### Traumatic Stress

The PTSD Checklist (PCL-5; Weathers et al., 2013) is a 20-item self-report scale that measures the DSM-5 symptoms of posttraumatic stress disorder (PTSD). Participants were asked to endorse PTSD symptoms on a 5-point Likert scale from 0 (n*ot at all*) to 4 (*extremely*). More specifically, participants were asked to respond to the prompt, “In the past month, how much were you bothered by …” High scores on PCL-5 are indicative of high acknowledgment of PTSD symptoms. Internal consistency for the PCL-5 from α = .56 to α = .95 (Moody & Lewis, 2019; Sveen et al., 2016). The internal reliability for the current study was α = .94. High test-retest reliability (r = .84) was reported for the PCL-5 (Bovin et al., 2016). Convergent validity was statistically significant with similar measures of trauma such as the PTSD Symptom Scale– Interview (r = .68), Trauma-Related Guilt Inventory-Hindsight Bias (r = .32), and Trauma-Related Guilt Inventory-Wrongdoing (r = .34) (Wortmann et al., 2016).

### Psychosocial Resources

Four psychosocial resources were tested as potential moderators of the association between GRMS and traumatic stress. First, to assess *resilience*, The Connor-Davidson Resilience Scale (CD-RISC; Connor & Davidson, 2003) is a 25-item measure that measures trait resilience. Sample items include “I am able to adapt when changes occur”, “I can deal with whatever comes my way”, and “I believe I can achieve my goals, even if there are obstacles.” Response options range from 1 (*not true at all*) to 5 (*true nearly all the time*). Higher scores are indicative of high levels of resilience. Good internal reliability was demonstrated with scores ranging from α = .91 to α = .93 (Brown & Tylka, 2011; Castelin & White, 2022). The internal reliability for the current study was high (α = .92).

Second, the Multidimensional Scale of Perceived Social Support (MSPSS; Zimet et al., 1988) is a 12-item measure that assesses perceived social support. The MSPSS evaluates support from three categories: friends, family, and significant others. Sample items include “There is a special person who is around when I am in need”, “My family really tries to help me”, and “I can count on my friends when things go wrong.” Response options ranged from 1 (*very strongly disagree*) to 7 (*very strongly agree*). Scores are averaged to produce a total score. Higher scores indicate perceptions of higher social support. Good internal reliability was demonstrated with scores ranging from α = .91 to α = .93 (Brown, 2008; Canty-Mitchell & Zimet, 2000). The internal reliability for the current study was α = .93. Discriminant validity was demonstrated with correlations between the Adolescent Family Caring Scale and the 3 MSPSS subscales (i.e., family, friend, significant other) (Canty-Mitchell & Zimet, 2000).

Third, *mastery* is a 7-item scale that assesses a person’s perceived control over one’s life (Pearlin & Schooler, 1978). Sample items include “There really is no way I can solve some of the problems I have” and “What happens to me in the future mostly depends on me.” Participants’ responses ranged on a Likert-type scale from 1 (*strongly agree*) to 7 (*strongly disagree*). Items are averaged to produce a score. A higher score reflects higher levels of mastery. The internal reliability of the scale is moderately high (α = .77) (Pearlin & Schooler, 1978). The internal reliability for the current study was moderately high (α = .77).

Last, the Rosenberg Self-Esteem Scale (RSE; Rosenberg, 1986) is a 10-item scale that assesses global self-worth. Sample items include “I feel that I am a failure” and “I feel that I have a number of good qualities.” Responses are endorsed on a Likert-type scale ranging from 1 (*strongly disagree)* to 4 (*strongly disagree*). Some items were reverse coded so that higher scores reflected higher self-esteem. Scores are totaled, and higher scores are indicative of higher self-esteem. High reliability is reported (α = .88) and high test-retest reliability with correlations ranging from (r = .85) to (r = .88) (Rosenberg, 1979). The internal reliability for the current study was (α = .77). Convergent validity was demonstrated with significant correlations to similar measures of self-esteem, such as the Coopersmith Self-Esteem Inventory (Rosenberg, 1979). As well, discriminant validity was demonstrated based on correlations between the Adolescent Family Caring Scale (AFCS) and the 3 MSPSS subscales (i.e., family, friend, significant other) (Canty-Mitchell & Zimet, 2000).

### Demographics

All analyses are adjusted for a first-generation college student (=1), sexual minority status (=1), age (18-22 years [reference], 23-30 years, 31-40 years, 41 years or older), work status (full-time, part-time, not working/other [reference]), and marital status (single [reference], married, other).

### Data Analysis

We first report descriptive statistics (mean, standard deviation, range) of all study measures. Next, we report correlations between key study measures in Table 3. Then, we performed multiple linear regression analysis to assess the association between GRMS overall, GRMS subscales, and traumatic stress (Table 3). To test for moderation, we performed statistical interactions between GRMS and each psychosocial resource (i.e., resilience, social support, mastery, and self-esteem) to assess whether psychosocial resources moderated the relationship between GRMS and traumatic stress. Table 4 includes the results of the moderation analysis. Figures 1-3 include visual displays of significant interactions.

**Figure 1:**
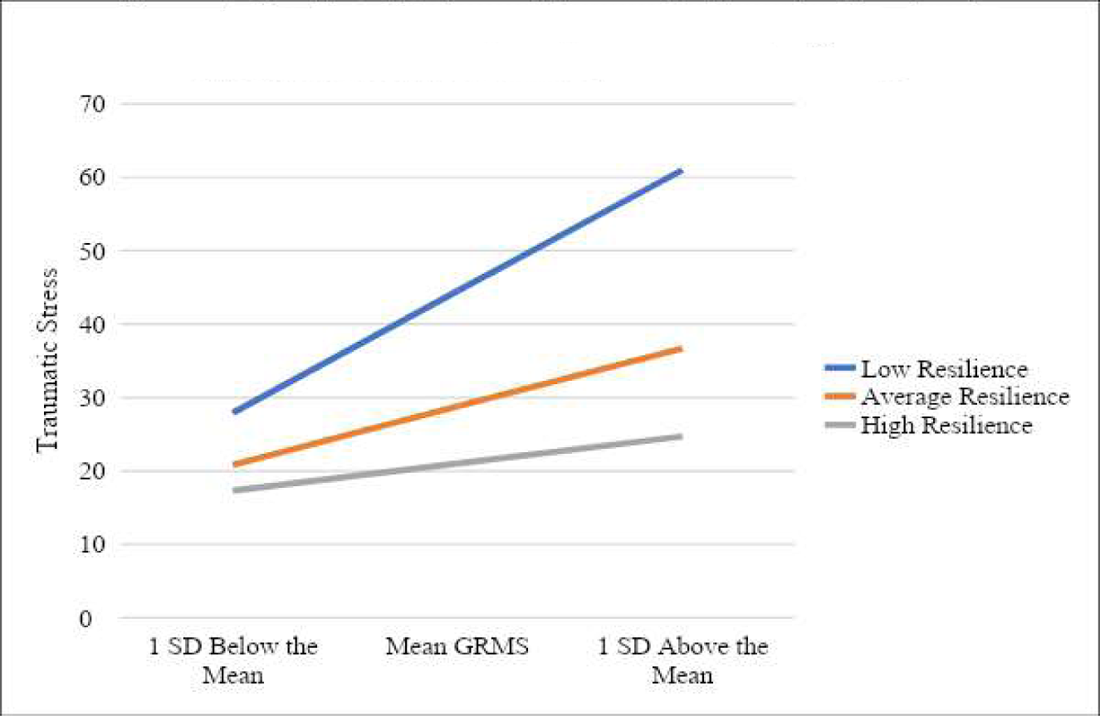
Predicted Values of Traumatic Stress by Gendered Racial Microaggressions (GRMS) and Resilience.

**Figure 2:**
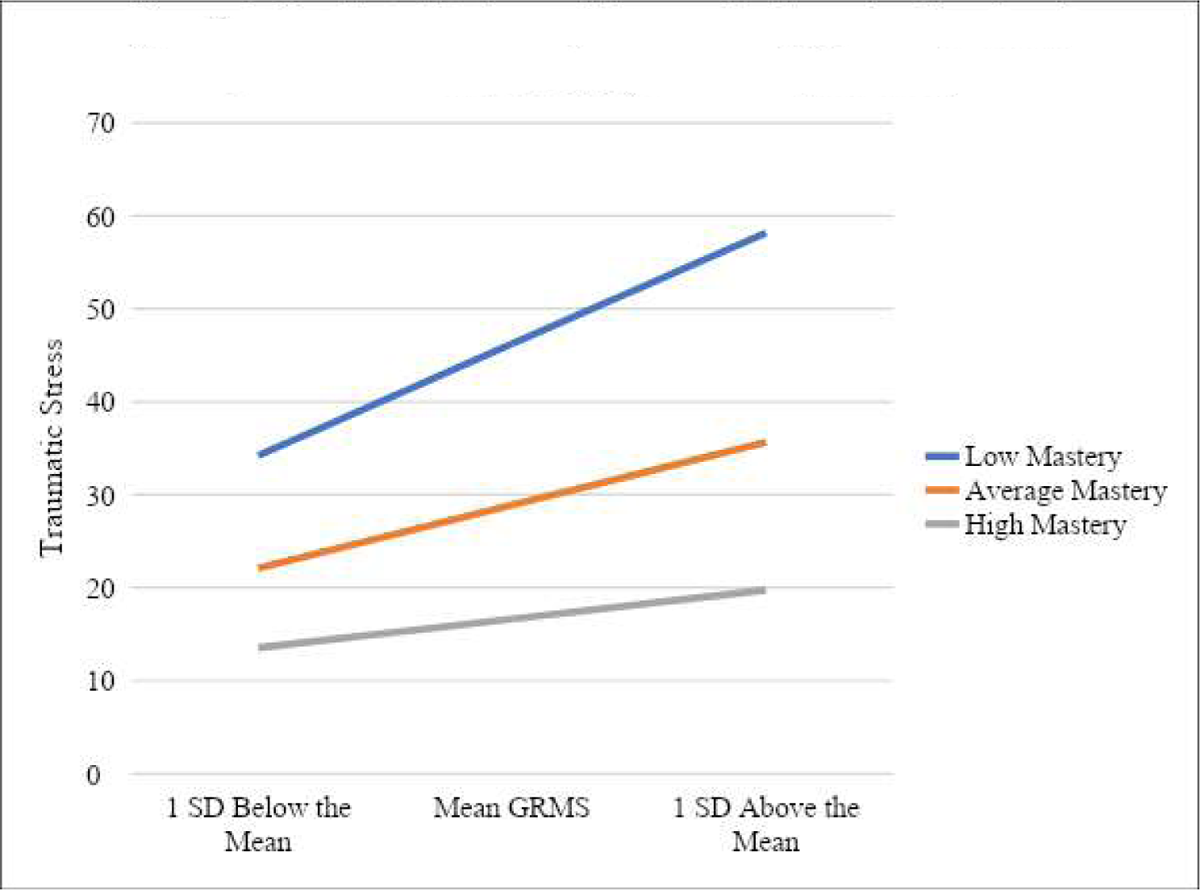
Predicted Values of Traumatic Stress by Gendered Racial Microaggressions (GRMS) and Mastery.

**Figure 3:**
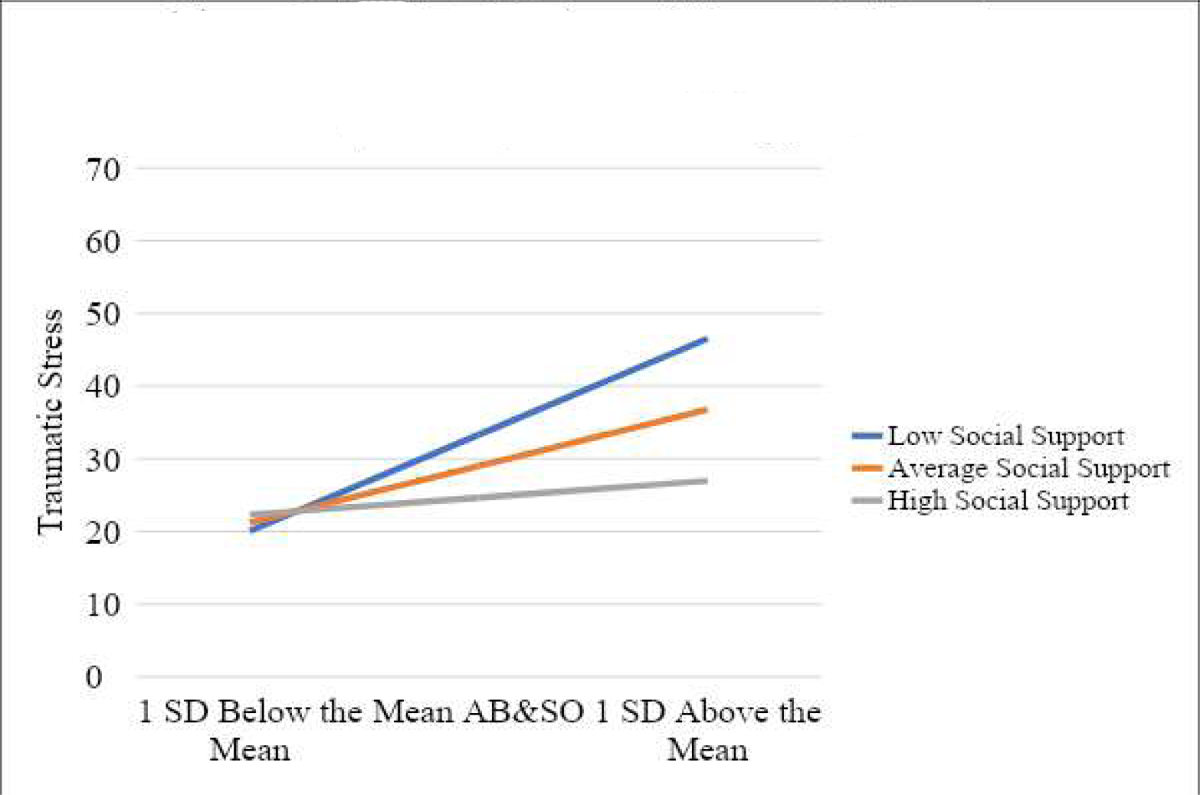
Predicted Values of Traumatic Stress by Assumptions of Beauty and Sexual Objectification (AB&SO) and Social Support.

## Results

### Preliminary Analyses

The survey included an initial sample of 234 respondents. Out of this sample, 33 respondents were excluded due to missing data. Specifically, respondents had missing data on traumatic stress (N = 11), GRMS (N = 5), resilience (N = 4), social support (N = 7), mastery (N = 12), and self-esteem (N = 8). With regards to sociodemographic data, respondents were missing on sexual orientation (N = 2), and age (N = 19). To retain as much of the sample as possible, we imputed age for the nineteen respondents missing data on age by using the mean age of respondents within their reported college standing category (e.g., sophomore). After only retaining respondents with complete cases on all other study measures, the restricted sample for this analysis is 201. All analyses were conducted with STATA 16.1 (StataCorp 2019).

See Table 1 for descriptive statistics for study measures. Pearson product-moment correlations indicated significant correlations between variables of interest (see Table 2); for instance, GRMS was positively associated with traumatic stress (r = .48). GRMS also significant and negatively correlated with resilience (r = −.18), social support (r = −.22), mastery (r = −30), and self-esteem (r = −.38). Psychosocial resources were negatively linked to traumatic stress.

**Table 1:** Means, Standard Deviations, and Range for All Study Variables (N = 201), Predicted values based on Model 1 from Table 4.

| | Mean/Prop. | SD | $\alpha$ | Min. | Max. |
| --- | --- | --- | --- | --- | --- |
| PTSD Symptom Checklist (PCL5-20) | 28.95 | 17.99 |  | .00 | 75.00 |
| Gendered Racial Microaggressions Scale (GRMS) | 1.88 | .97 | .95 | .00 | 4.00 |
| GRMS Subscales |  |  |  |  |  |
| Assumptions of Beauty and Sexual Objectification | 1.69 | 1.06 |  | .00 | 4.00 |
| Silenced and Marginalized | 1.91 | 1.12 |  | .00 | 4.00 |
| Strong Black Woman Stereotype | 2.00 | 1.05 |  | .00 | 4.00 |
| Angry Black Woman Stereotype | 2.31 | 1.25 |  | .00 | 4.00 |
| Psychosocial Resources |  |  |  |  |  |
| Connor-Davidson Resilience Scale | 3.95 | .57 | .92 | 1.79 | 5.00 |
| Multidimensional Scale of Perceived Social Support | 5.39 | 1.19 | .93 | 1.25 | 7.00 |
| Pearlin Mastery Scale | 4.98 | 1.06 | .77 | 3.00 | 7.00 |
| Rosenberg Self-Esteem Scale | 3.06 | .61 | .88 | 1.10 | 4.00 |
| **Control Measures** |  |  |  |  |  |
| First-Generation College Student | .39 |  |  |  |  |
| Sexual Minority | .16 |  |  |  |  |
| Age |  |  |  |  |  |
| 18-22 years (reference) | .60 |  |  |  |  |
| 23-30 years | .17 |  |  |  |  |
| 31-40 years | .09 |  |  |  |  |
| 41 years + | .13 |  |  |  |  |
| Work Status |  |  |  |  |  |
| Full-time | .35 |  |  |  |  |
| Part-time | .37 |  |  |  |  |
| Not Working/Other (reference) | .28 |  |  |  |  |
| Marital Status |  |  |  |  |  |
| Single (reference) | .76 |  |  |  |  |
| Married | .14 |  |  |  |  |
| Other | .10 |  |  |  |  |

Source: Gendered Racism and Well-Being Questionnaire (2020-2021)

**Table 2:** Bivariate Pearson’s Correlations for Key Dependent and Independent Measures (N = 201), Predicted values based on Model 3 of Table 4.

|  | 1. | 2. | 3. | 4. | 5. | 6. | 7. | 8. | 9. |
| --- | --- | --- | --- | --- | --- | --- | --- | --- | --- |
| 1. PTSD Symptom Checklist (PCL5-20) | 1 |  |  |  |  |  |  |  |  |
| 2. GRMS | .48* | 1 |  |  |  |  |  |  |  |
| 3. AB & SO | .52* | .93* | 1 |  |  |  |  |  |  |
| 4. Silenced and Marginalized | .38* | .89* | .75* | 1 |  |  |  |  |  |
| 5. Strong Black Woman Stereotype | .28* | .81* | .64* | .64* | 1 |  |  |  |  |
| 6. Angry Black Woman Stereotype | .45* | .80* | .66* | .65* | .67* | 1 |  |  |  |
| 7. Connor-Davidson Resilience Scale | -.34* | -.18* | -.19* | -.19* | -.01 | -.20* | 1 |  |  |
| 8. Social Support | -.37* | -.22* | -.19* | -.23* | -.13 | -.23* | .55* | 1 |  |
| 9. Pearlin Mastery Scale | -.50* | -.30* | -.32* | -.27* | -.11 | -.28* | .58* | .46* | 1 |
| 10. Rosenberg Self-Esteem Scale | -.51* | -.38* | -.30* | -.29* | -.08 | -.28* | .64* | .52* | .67* |

Source: Gendered Racism and Well-Being Questionnaire (2020-2021)
GRMS = Gendered Racial Microaggressions Scale

In terms of GRMS subscales, each GRMS dimension was positively associated with traumatic stress, with the largest correlations observed for Assumptions of Beauty & Sexual Objectification (r = .52) followed by Angry Black Woman Stereotype (r = .45), Silenced & Marginalized (r = .38) and Strong Black Woman Stereotype (r = .28). Three dimensions of GRMS were negatively associated with resilience, social support, mastery, and self-esteem. The one exception was the Strong Black Woman Stereotype, which was not significantly correlated with any psychosocial resources.

### Multiple Regression Analysis: Direct Associations

In Table 3, the association between GRMS and traumatic stress was assessed. In Model 1, GRMS was associated with increased traumatic stress symptoms (β = 8.73, SE = 1.14, p <.001), and along with controls, explained 33% of the variation in traumatic stress (R2 = .33). Models 2-5 assessed the association between each GRMS dimension and traumatic stress. Each GRMS subscale was positively associated with traumatic stress. Specifically, Assumptions of Beauty and Sexual Objectification (Model 2; β = 8.24, SE = 1.04, p <.001), Silenced and Marginalized (Model 3; β = 6.13, SE = 1.03, p <.001), Strong Black Woman (Model 4; β = 5.37, SE = 1.12, p <.001), and Angry Black Woman (Model 5; β = 6.11, SE = .92, p <.001) were associated with higher traumatic stress. In Model 6, all four GRMS dimensions were included in the same model and two subscales remained positively associated with traumatic stress: Assumptions of Beauty and Sexual Objectification (β = 7.09, SE = 1.71, p <.001) and Angry Black Woman (β = 3.64, SE = 1.14, p <.01). Overall, accounting for controls and four GRMS dimensions explained 37% of the variation in traumatic stress symptoms (R-squared = .37).

**Table 3:** OLS Regression of the Association between Gendered Racial Microaggressions Scale (GRMS) Overall, GRMS Subscales, and Traumatic Stress (N = 201)

|  | Model 1 | Model 2 | Model 3 | Model 4 | Model 5 | Model 6 |
| --- | --- | --- | --- | --- | --- | --- |
| **GRMS Overall** | 8.73***<br>(1.14) |  |  |  |  |  |
| **GRMS Subscales** |  |  |  |  |  |  |
| AB & SO |  | 8.24***<br>(1.04) |  |  |  | 7.09***<br>(1.71) |
| Silenced and Marginalized |  |  | 6.13***<br>(1.03) |  |  | -.19<br>(1.58) |
| Strong Black Woman |  |  |  | 5.37***<br>(1.12) |  | -2.14<br>(1.57) |
| Angry Black Woman |  |  |  |  | 6.11***<br>(.92) | 3.64**<br>(1.32) |
| Constant | 12.96***<br>(3.31) | 14.69***<br>(3.10) | 18.75***<br>(3.20) | 19.91***<br>(3.44) | 15.90***<br>(3.31) | 12.78***<br>(3.31) |
| R<sup>2</sup> | .33 | .34 | .26 | .22 | .29 | .37 |

Source: Gendered Racism and Well-Being Questionnaire (2020-2021)
Standard errors in parentheses.
GRMS = Gendered Racial Microaggressions Scale
AB & SO = Assumptions of Beauty and Sexual Objectification
All models adjusted for first-generation college student status, age, sexual identity, work status, and marital status.
\* $p < 0.05$ , \*\* $p < 0.01$ , \*\*\* $p < 0.001$

### Multiple Regression Analysis: Moderation

The next aim of the study was to assess whether psychosocial resources moderated the association between GRMS and traumatic stress. See Table 4 for the moderation analysis. Evidence of moderation was identified for two psychosocial resources: resilience (see Model 1) and mastery (Model 3). No moderation effects for social support or self-esteem were observed.

**Table 4:**
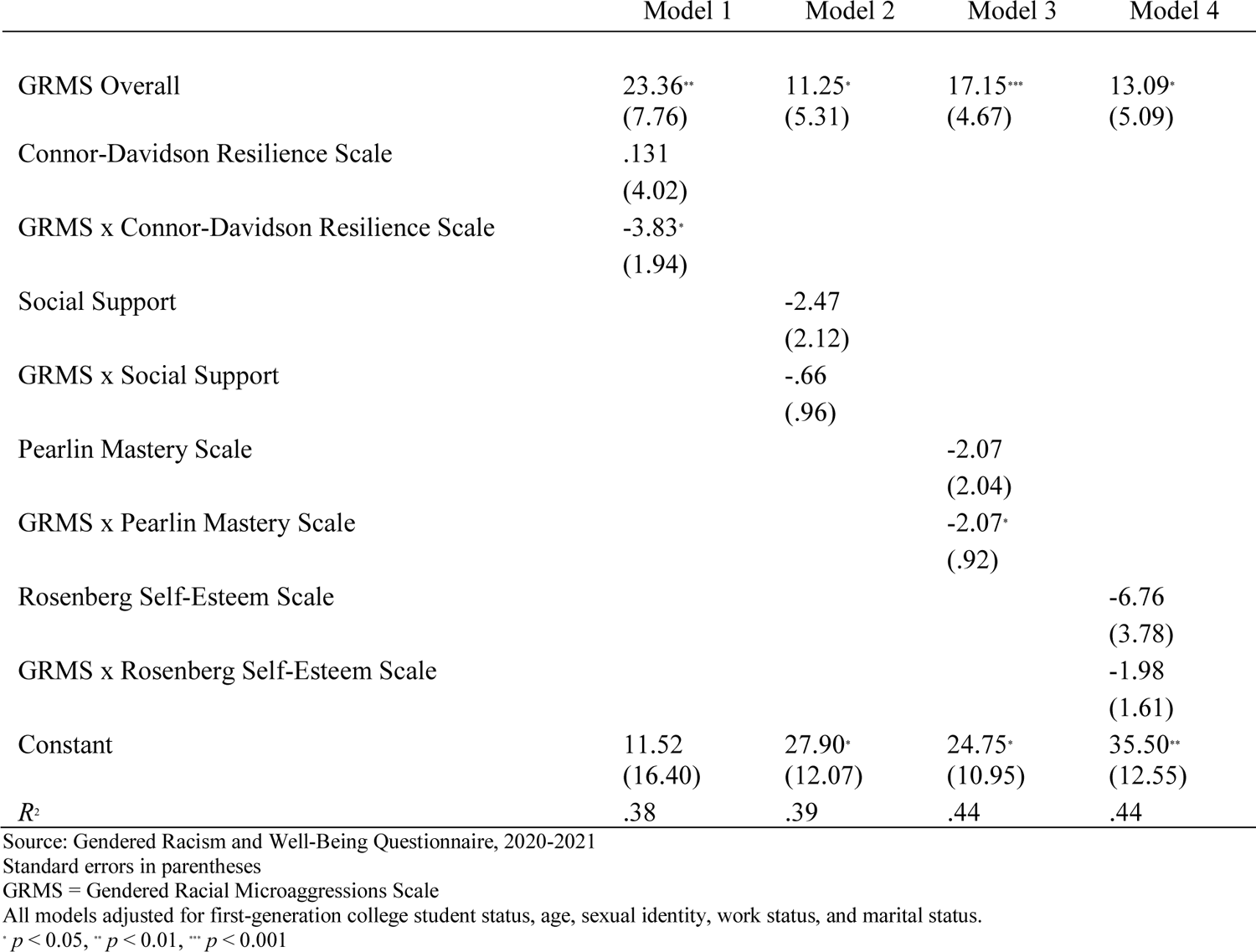
OLS Regression of the Moderating Effects of Psychological Resources in the GRMS-Traumatic Stress Association (N = 201), Predicted values based on an interaction between AB&SO and social support, adjusting for study controls (model available upon request).

Figure 1 displays the moderating role of resilience. Predicted values of traumatic stress symptoms by GRMS and resilience are shown. All other measures were set at their means. Values of resilience and GRMS were assessed as low (1 SD below the mean), average (mean), and high (1 SD above the mean) levels. While the association between GRMS and traumatic stress is strongest among individuals with low resilience, high resilience is protective (i.e., associated with lower traumatic stress) even when GRMS levels are high.

In Figure 2, the moderating effect of mastery is shown. A similar approach to predicted values is used as explained above. For individuals with low mastery, there is a strong and positive association between GRMS and traumatic stress. However, this association is less pronounced for those with average and high mastery levels.

When moderation was examined for GRMS subscales, Assumptions of Beauty and Sexual Objectification were moderated by resilience and mastery, suggesting that this component of GRMS is moderated by these two factors. Social support moderated the association between Assumptions of Beauty and Sexual Objectification and traumatic stress symptoms, as shown in Figure 3. That is, when social support is low, the association between Assumptions of Beauty and Sexual Objectification and traumatic stress is strong and positive. This positive association is less pronounced among those with average social support, and individuals with high social support are protected against the traumatic stress effects.

## Discussion

This study examined the association between Black women’s experience of gendered racial microaggressions (GRMS) and traumatic stress. To date, there are only two other studies that investigated Black women’s experience of GRMS, racial socialization, and traumatic stress (Moody & Lewis, 2019) and GRMS, racial discrimination, HIV-related discrimination, and posttraumatic stress cognitions and symptoms (Dale & Safren, 2019). Building on these two studies, we evaluated the different types of GRMS and their association with traumatic stress. We evaluated the moderating effects of psychosocial resources, specifically, resilience, social support, mastery, and self-esteem, on the relationship between GRMS and traumatic stress.

Consistent with previous studies, the findings demonstrate that heightened experiences of GRMS contribute to increased symptoms of traumatic stress (Dale & Safren, 2019; Moody & Lewis, 2019). And thus, our first hypothesis that greater endorsement of GRMS will positively relate to traumatic stress was supported. As Black women experience a greater frequency of GRMS, it is likely that potential symptoms of traumatic stress will increase. The process of healing for Black women must begin with helping them understand their experiences of gendered racism, empowering them to recognize traumatic stress symptoms, and cultivating critical consciousness about the impacts of intersecting systemic oppressions (Comas-Díaz, 2016; Quiñones-Rosado, 2020).

Individually, the GRMS subscales, Assumptions of Beauty & Sexual Objectification, Angry Black Woman Stereotype, Silenced & Marginalized, and Strong Black Woman Stereotype, statistically and positively related to traumatic stress. These findings suggest that experiencing any type of gendered racial microaggression will contribute to higher levels of traumatic stress. Assumptions of Beauty and Sexual Objectification had the strongest connection with traumatic stress. Thus, socio-cultural or political factors such as images in social media, racialized stereotypes (e.g., Jezebel) perpetuated in music videos or television shows, Eurocentric standards of beauty (e.g., thinness, lily complex, curviness), or any other systemic oppressive factors likely contribute to Black women’s vulnerability to internalization of negative messaging (e.g., insecure and undervalued) and susceptibility trauma or other mental health conditions (Berger, 2018; Cheers, 2020; Jones & Shorter-Gooden, 2003; Lewis et al., 2016).

The second hypothesis about increased reported experiences of GRMS will negatively relate to psychosocial resources (resilience, social support, mastery, and self-esteem) was partially supported. Black women who endorsed a higher frequency of GRMS likely experience lower levels of psychosocial resources. All the GRMS subscales, except the Strong Black Women stereotype, were associated with fewer psychosocial resources. This finding is unsurprising, as the Strong Black woman stereotype did not relate significantly to resilience in another study as well (Castelin & White, 2022).

Our third hypothesis that psychosocial resources would be inversely associated with traumatic stress was supported. Consistent with previous research, psychosocial resources can play a role in reducing levels of traumatic stress (Dale & Safren, 2019; Watson et al., 2016; Zanbar et al., 2021). It is essential that we continue to investigate additional strengths (e.g., spirituality or mindfulness) and self-care strategies (e.g., yoga) to facilitate harmony and balance (Berger, 2018; Patton & McClure, 2009; Wright & Lewis, 2020).

Our final hypothesis that the psychosocial resources would reduce the effects of the relationship between GRMS and traumatic stress was partially supported. We offer evidence that psychosocial resources can serve as a buffer for Black women experiencing gendered racism (Lewis et al., 2012; Neal-Barnett et al., 2011; Zanbar et al., 2021). Our results revealed that resilience and mastery serve as protective factors, reducing the contributions of GRMS to traumatic stress. This finding mirrors a previous study that demonstrates Black women favoring the Strong Black women stereotype and who have high levels of resilience, experience reduced mental health symptoms and decreased suicidal behaviors (Casteline & White, 2022). A focus on factors that facilitate resilience development can be a step in Black women’s healing (Brown, 2008; Mushonga et al., 2021). Complementary to previous research, we present evidence that the Black women who reported at least average levels of mastery had a reduced association between GRMS and traumatic stress. This is the first study to find that self-mastery reduces the effects of GRMS on traumatic stress for Black women. Mastery is a factor that can complement the presence of resilience for Black women, reducing the effects of stress and susceptibility to psychological distress (Erving et al., 2021; Glister, 2016; Keith et al., 2010). Lastly, the beauty and objectification microaggressions were moderated by resilience and mastery, suggesting a less profound effect of traumatic symptoms.

Our findings show that social support has some effect on the relationship between Assumptions of Beauty and Sexual Objectification and traumatic stress. In other words, some Black women with average or high social support can have less profound symptoms of traumatic stress in response to experiencing microaggressions related to beauty and sexual objectification. However, those with low social support may be more vulnerable to the full effect of traumatic stress. While social support appears to serve as a protective factor for Black women experiencing microaggressions related to beauty and objectification, it does not seem to have any effect on the other microaggressions. This finding is inconsistent with existing literature (e.g., Erving et al. 2022). Previous researchers advocate for the need for social support, informal or formal, for Black women as it has an impact on reducing psychological distress (Brown, 2008; Constantine et al., 2002; Neal-Barnett et al., 2011). Perhaps the MSPSS failed to capture the type of social support that the Black women in this sample experience.

Unlike social support, self-esteem did not have any effect on the relationship between GRMS and traumatic stress. It is possible that self-esteem could inform extraneous variables that have the actual effect on the relationship between GRMS and traumatic stress. For example, self-esteem is a construct that informs gendered racial socialization (Brown et al., 2017), racialized body dissatisfaction (Winter et al., 2019), and internalized racial oppression (Bailey et al., 2011), which could be impactful as opposed to self-esteem alone. As psychosocial resources appear to serve as protective factors, it is likely there are also some that serve as risk factors that may have a null or opposite effect (Moody & Lewis, 2019).

### Practice Implications

The findings of our study offer several implications for clinical practice. First, it is necessary for health service providers to be mindful of the ways in which existing disparities exacerbate Black women’s psychological distress. Second, mirroring previous studies, our findings highlight the extent and degree to which Black women experience GRMS. When conceptualizing Black women’s presenting concerns and symptoms, clinicians need to account for types of microaggressions experienced and susceptibility to traumatic stress. Third, non-Black practitioners must consider the detrimental effects of gendered racism and screen for these experiences in the diagnostic evaluation. Facilitating a dialogue about Black women’s racial and gendered experiences and the connection with trauma-related symptoms in addition to typical diagnostic screening data is essential. Fourth, practitioners must exercise cultural humility and sensitivity as well as be comfortable with providing safe, affirming, and nonjudgment space for exploration of racial, gendered, or other cultural narratives. Lastly, the most notable of the findings were the effects of resilience and mastery on the relationship between GRMS and traumatic stress.

### Limitations and Future Directions

Caution should be exercised when generalizing the findings given that the results are reflective of a homogenous population. The sample consisted of undergraduate and graduate women at an HBCU. Investigating the hypotheses with a nationally representative sample could offer generalizable findings. Some of the coefficients did not reach statistical significance; thus, replicating the study with a larger sample size could strengthen or alter the results identified here. Social support was less impactful in the gendered racism and traumatic stress association than previously anticipated. Future studies should include measures that assess a broader range of social support. Self-esteem did not act as a moderator, and thus, perhaps it plays more of an indirect role in the relationship between gendered racism and traumatic stress. Further investigation identifying risk factors and effects on Black women’s vulnerability to traumatic stress resulting from gendered racism is needed.

## Conclusion

Given Black women’s elevated risk of PTSD compared to other women, it is critical to identify risk factors within this population. Our study revealed a significant contribution of GRMS, a form of intersectional stress, contributing to traumatic stress among college-age Black women. Microaggressions about Assumptions of Beauty and Sexual Objectivation and the Angry Black Woman stereotype were particularly harmful. Black women’s individual strengths, particularly resilience and mastery, reduced the psychological harm of GRMS; social support from family, friends, and a “significant other” collectively shielded Black women psychologically from certain kinds of microaggressions. Lastly, using a strengths-based treatment approach can highlight strengths in clients that might serve as protective factors.

## Compliance with Ethical Standards Disclosure of Potential Conflicts of Interest

There are no conflicts of interest pertaining to the authors to report or special circumstances for this work.

## Declaration of Funding

This manuscript and the work completed in preparation for this manuscript were supported by funding awarded to Tiffany R. Williams from the American Association of University Women for Postdoctoral Research Leave. No other funding sources, grants, or financial agreements were used.

## Research involving Human Participants and/or Animals

The research included human participants. No animals were included in the research. The project was approved by the Tennessee State University Institutional Review Board (##HS-2020-4487) before data collection. The authors followed the American Psychological Association’s ethical guidelines throughout the research and reporting process. The authors also received CITI research training prior to the collection of data.

## Informed Consent

The first page of the online survey included a document with the following information: a brief summary of the research, purpose, procedures, risks, benefits/compensation, confidentiality, and rights to withdraw at any time provided prior to participation in the study. Participants provided their consent before moving forward to the study instruments in the online survey. The authors also utilized the American Psychological Association’s ethical guidelines throughout the research and writing process.

## Data Availability

All data produced in the present study are available upon reasonable request to the authors.

## Notes

### Competing Interest Statement

The authors have declared no competing interest.

### Author Declarations

The Institutional Review Board (ethics committee) of Tennessee State University (HS-2020-4487) gave ethical approval for this work.

